# Sub-Analysis of a Randomized Controlled Trial of Neuromuscular Electrostimulation of the Common Peroneal Nerve after Forefoot Surgery

**DOI:** 10.64898/2026.08.21.26361007

**Authors:** Anne-Marie Piftor, Duncan Shirreffs Bain, Kieron Day

## Abstract

Gaps remain in the evidence base for postoperative management following forefoot surgery. A recent randomized controlled trial (ClinicalTrials.gov NCT04927234) demonstrated improved outcomes with intermittent one Hertz (Hz) neuromuscular electrical stimulation (NMES) of the common peroneal nerve. This sub-analysis evaluates its effect in patients undergoing forefoot surgery.

Forty-two patients undergoing forefoot procedures were included; 26 received NMES plus standard of care (SOC) and 16 received SOC alone. Wound healing was assessed at 14 days. Edema was measured using the figure-of-eight (FO8) method. Patient-reported outcomes were assessed using the Manchester-Oxford Foot Questionnaire (MOXFQ).

At 14 days, complete wound healing occurred in 77% of patients receiving NMES plus SOC compared with 40% in the SOC group (p<0.05). Edema reduction was significantly greater in the NMES group, with a 74% relative reduction compared with SOC (p=0.02).

Intermittent one Hz NMES of the common peroneal nerve was associated with improved wound healing and reduced postoperative edema following forefoot surgery.

## Introduction

Hallux valgus correction is among the most commonly performed foot procedures, reflecting the high prevalence of the condition across adult populations.^1–4^ Despite the wide range of surgical techniques for hallux valgus correction described,^5^ postoperative recovery remains variable, and recurrence has been associated with factors such as non-compliance with postoperative instructions.^6^

Clinical consensus statements have highlighted variability in treatment pathways and identified gaps in the evidence base, particularly in relation to postoperative care, thromboprophylaxis, outcome measurement, and rehabilitation.^7, 8^ Patient-reported outcome measures may also have limitations in reliability, validity, and responsiveness, reducing their ability to discriminate between treatment strategies.^9, 10^ Postoperative swelling is common following forefoot surgery^11^ and represents a key target for postoperative management, as it may impact wound healing, mobility, and recovery.^12, 13^

A recent randomized controlled trial^14^ demonstrated that intermittent one Hertz neuromuscular electrical stimulation (NMES) of the common peroneal nerve reduces postoperative edema by 33% (p<0.05) in patients undergoing foot and ankle surgery (ClinicalTrials.gov: NCT04927234). This intervention delivers a painless transdermal stimulus that activates the venous muscle pump.^15^ A substantial proportion of patients in that study underwent forefoot procedures. The aim of the present analysis was to evaluate whether this intervention is similarly effective within this subgroup.

## Methods

This study represents a sub-analysis of a larger open-label, multicenter, prospective randomized controlled trial of patients undergoing foot and ankle surgery.^14^ Participants were recruited from international participating centres in the United States, Spain, and the United Kingdom and were scheduled to undergo elective forefoot surgery. Eligible procedures included hallux valgus correction, Weil osteotomy, lesser toe surgery, first metatarsophalangeal joint (MTPJ) cheilectomy, first MTPJ fusion, and forefoot reconstruction.

Participants were randomized in a 1:1 ratio to receive standard of care (SOC) plus neuromuscular electrical stimulation (NMES) or SOC alone. Patients and the public were not involved in the design, conduct, or reporting of the study.

### Ethical Approval and Consent

Ethical approval for the parent randomized controlled trial was obtained from the relevant ethics committees in the United Kingdom, Spain, and the United States: West of Scotland REC 4 (21/WS/0099), CEIm Hospital Clínic de Barcelona (HCB/2021/1251), CEIm Hospital Universitari Vall d’Hebron (PR(AT)392/2022), and the Western Institutional Review Board/WCG (20221183).

Written informed consent was obtained from all participants prior to enrolment. The trial was registered on ClinicalTrials.gov (NCT04927234) and conducted in accordance with the Declaration of Helsinki, EN ISO 14155:2020, and ICH E6(R2) Good Clinical Practice.

### Eligibility Criteria

*Inclusion Criteria:* age ≥18 years; no more than two falls in the preceding 12 months; intact, healthy skin at the intended NMES application site; scheduled for forefoot and/or hindfoot surgery; ability to understand study requirements and provide written informed consent.

*Exclusion Criteria:* pregnancy; use of another neuromodulation device; lower limb trauma preventing NMES stimulation of the common peroneal nerve; absence of a motor response to NMES (no visible rhythmic dorsiflexion); or participation in another clinical study that could interfere with outcomes.

### Sample Size Determination

Sample size for the original study was based on the mean and standard deviation differences between study arms in previous studies.^16^ It was determined that 50 patients would be needed in each group for a power of 95% and type I error (alpha) of 0.05. The calculation was based on the formula:

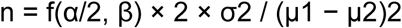

where μ1 and μ2 represent the mean outcomes in the SOC and SOC + NMES groups respectively, and σ is the standard deviation. Assuming an attrition rate of 18% of patients, 61 patients were planned for recruitment into each arm of the trial.

The sample size in this sub-analysis was smaller than originally calculated for the parent study due to attrition and missing outcome data, resulting in reduced statistical power.

### Randomization and Treatment

Participants were randomized in a 1:1 ratio using a centrally generated computer sequence via the Castor Electronic Data Capture platform to receive standard of care (SOC) plus neuromuscular electrical stimulation (NMES) or SOC alone, ensuring allocation concealment across all participating centres.

For this sub-analysis, only participants undergoing elective forefoot surgery were included. Participants in the SOC + NMES group received NMES in addition to SOC, while those in the SOC group received SOC alone. Standard of care consisted of non-steroidal anti-inflammatory drugs, limb elevation, and off-loading.

### Endpoints

The primary endpoint was postoperative edema. Secondary endpoints included the incidence of adverse events, postoperative pain measured using a visual analogue scale (VAS), surgical wound healing at 14 days postoperatively, and self-reported health status assessed using the Manchester-Oxford Foot Questionnaire (MOXFQ).

Edema was quantified using the validated figure-of-eight (FO8) method, which provides a standardized measure of foot and ankle swelling.^17^ The percentage difference between the involved and uninvolved limbs was calculated. FO8 measurements have demonstrated correlation with water displacement methods^18, 19^ and optical techniques,^20^ as well as good reliability (intraclass correlation coefficient 0.94) and validity (r=0.65, p<0.001).^21–23^

Measurements were performed with participants in a standardized supine position using predefined anatomical landmarks (Figure 1). Each measurement was repeated three times on both limbs and averaged. Assessments were performed by trained clinical staff who were not blinded to treatment allocation, using standardized techniques to minimise interobserver variability.

**Figure 1.**
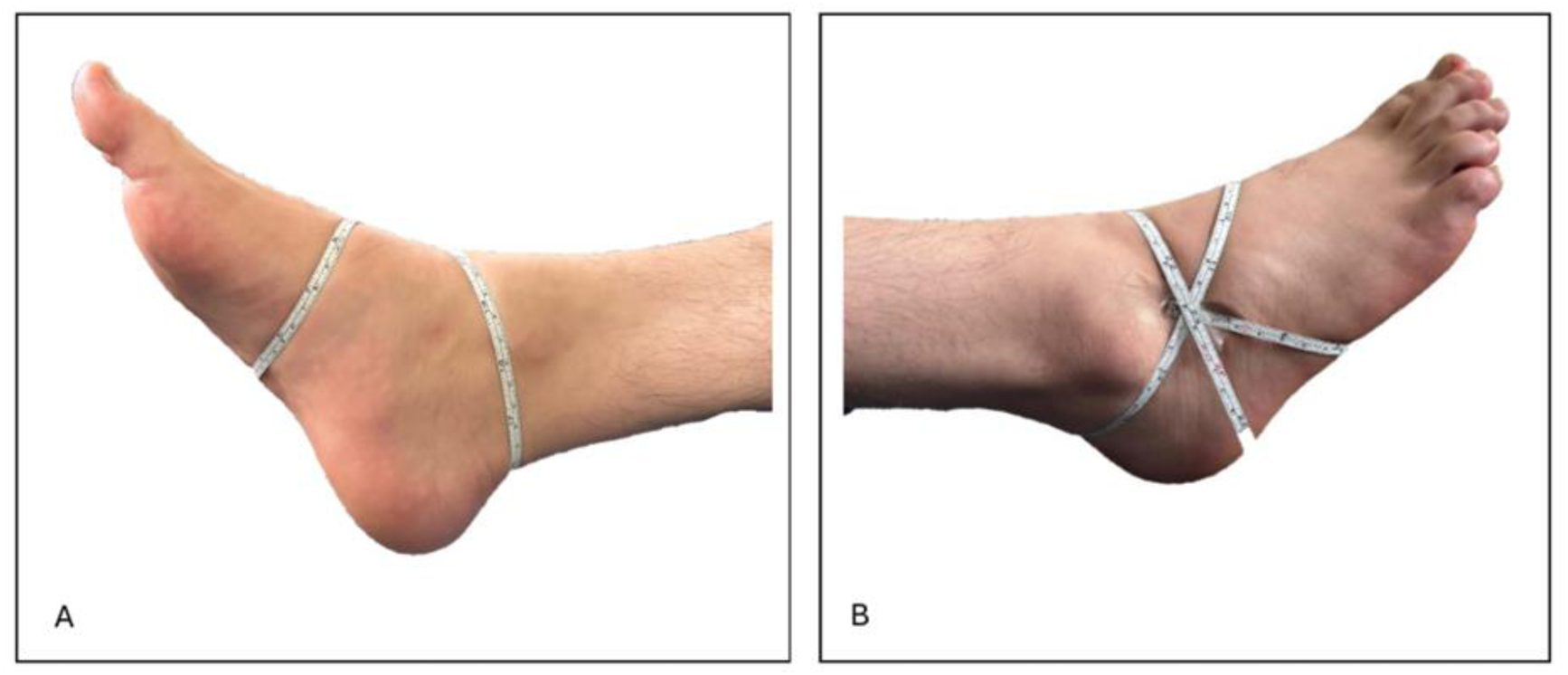
Figure-of-eight (FO8) method for assessment of foot and ankle edema. (A) Medial view and (B) lateral view demonstrating the standardized tape path applied across predefined anatomical landmarks.

### Intervention

The NMES device (geko® T device, Firstkind Ltd, Daresbury, Figure 2) has 11 stimulation settings designed to activate the venous muscle pumps of the calf and foot, irrespective of individual skin impedance. The optimal setting was defined as the lowest setting sufficient to produce an intermittent (one Hz) contraction of the leg muscles, observed as visible dorsiflexion/eversion twitch of the foot, while remaining comfortable for the patient.

**Figure 2.**
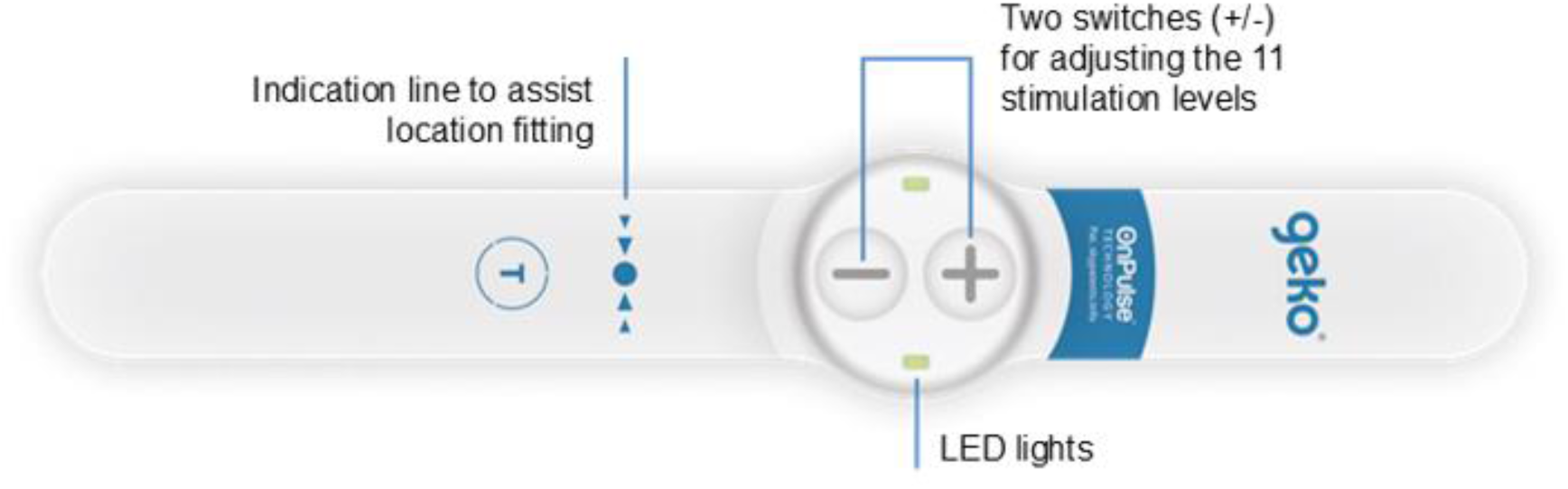
The geko® T device, Firstkind Ltd, Daresbury

The device was applied to the operated limb only, positioned just below the knee over the fibular head in accordance with the manufacturer’s instructions. NMES was delivered continuously for the first 24 hours postoperatively, followed by 12 hours per day at home until the day 14 (±2) follow-up visit.

### Statistical Analysis

Outcomes were compared between the SOC + NMES and SOC groups. Continuous variables were analysed using unpaired Student’s t-tests, and categorical variables using Fisher’s exact test. Results are presented with p-values and 95% confidence intervals where appropriate, with p<0.05 considered statistically significant.

Analyses were conducted using a modified intention-to-treat approach, including all randomized participants with available primary outcome data at the 14-day follow-up and analysed according to their allocated group. Missing data were not imputed.

This study represents a post hoc subgroup analysis of participants undergoing forefoot surgery from the parent randomized controlled trial. Descriptive analyses of baseline demographics, comorbidities, and surgical procedures were performed. No formal subgroup or sensitivity analyses were undertaken.

## Results

A total of 55 participants were screened and completed baseline assessment. All participants were randomized in a one:one ratio to receive either standard of care plus neuromuscular electrical stimulation (SOC + NMES; n = 31) or standard of care alone (SOC only; n = 24).

Following randomization, participants were excluded from the modified intention-to-treat analysis due to protocol non-compliance, missed or out-of-window followup visits, withdrawal of consent, or missing primary outcome data. At day 14, the modified intention-to-treat population comprised 26 participants in the SOC + NMES group and 16 participants in the SOC only group. Participant flow and reasons for exclusion are presented in Figure 3.

**Figure 3.**
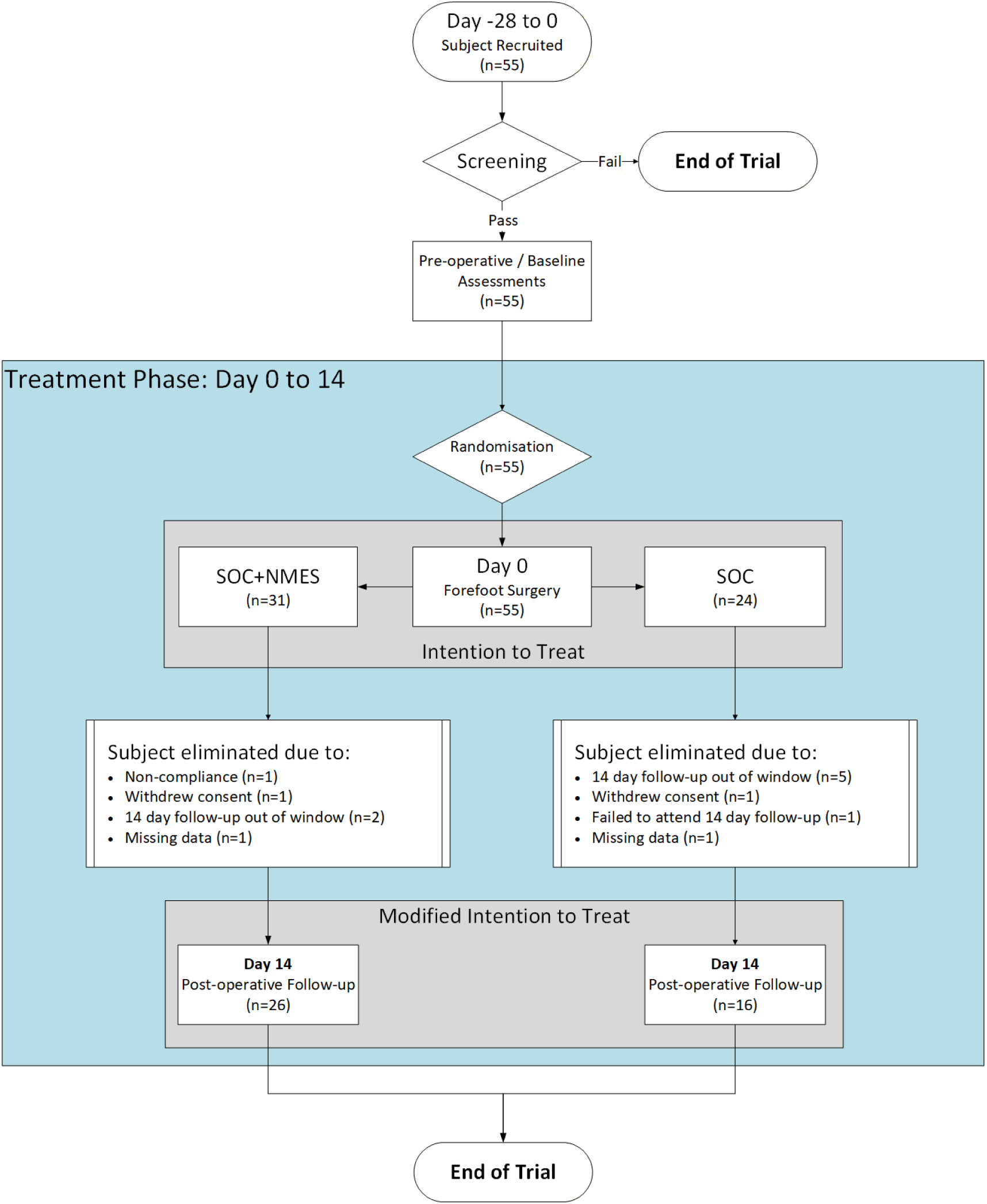
Participant flow diagram for the forefoot surgery sub-analysis. A total of 55 participants were randomized to receive standard of care plus neuromuscular electrical stimulation (SOC + NMES; n = 31) or standard of care alone (SOC only; n = 24). Participants were excluded from the modified intention-to-treat population due to protocol non-compliance, missed or out-of-window followup visits, withdrawal of consent, or missing primary outcome data. The final modified intention-to-treat population at day 14 comprised 26 participants in the SOC + NMES group and 16 participants in the SOC-only group.

### Baseline Demographic Characteristics

Table 1 presents the demographic characteristics of the randomized cohort (SOC, n = 24; SOC + NMES, n = 31), comparing the SOC + NMES group with the SOC group and reporting the mean and standard error (SE) for each parameter. In accordance with EU data protection regulations, demographic data were restricted to three identifiable parameters to prevent patient identification: age, body mass index (BMI), and sex.

**Table 1.** Baseline demographic characteristics of the study population. Values are presented as mean (standard error [SE]) unless otherwise indicated. Categorical variables are presented as number (percentage). Between-group comparisons for continuous variables were performed using Student’s *t*-test, and categorical variables were compared using Fisher’s exact test.

| Characteristic | SOC + NMES (n=31) | SOC (n=24) | p value |
| --- | --- | --- | --- |
| Age (years) | 58.67 (1.81) | 59.68 (2.75) | 0.75 |
| BMI | 27.04 (0.65) | 27.02 (0.86) | 0.98 |
| Males, n (%) | 4 (12.9%) | 7 (29.2%) |  |
| Females, n (%) | 26 (83.9%) | 18 (75.0%) |  |
| Not reported | 1 (3.2%) | 0 (0%) |  |

The groups were similar with respect to age and BMI, with no statistically significant differences observed between groups using Student’s t-test. The distribution of sex was also comparable between groups, with no statistically significant differences identified using Fisher’s exact test.

### Comorbidities

Baseline comorbidity characteristics of the randomized cohort are summarized in Table 2. Comorbidities were distributed across multiple system categories in both groups (SOC, n = 24; SOC + NMES, n = 31).

**Table 2.**
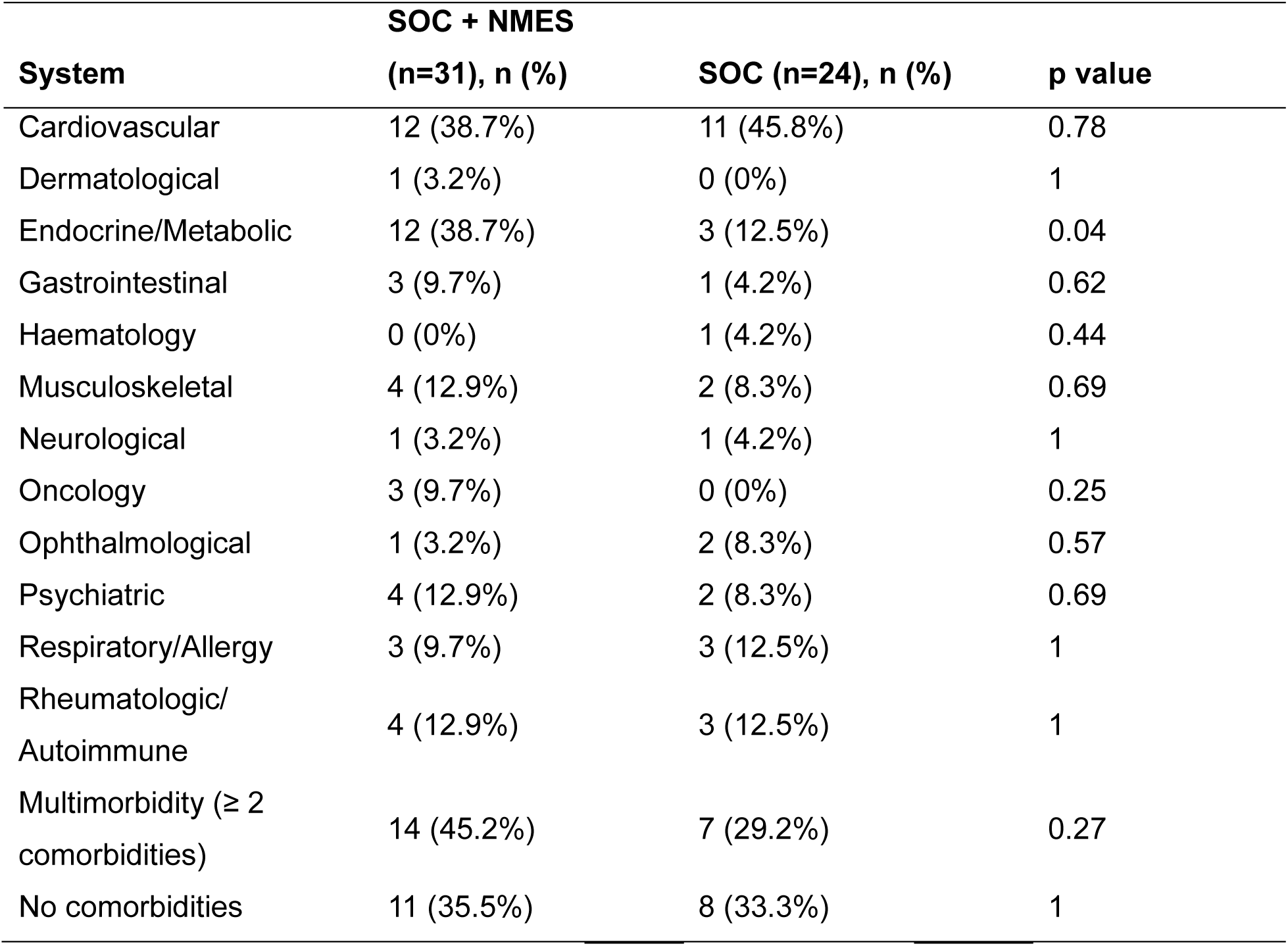
Distribution of comorbidities by system category in the intervention (n = 31) and standard of care (SoC, n = 24) groups.

The overall prevalence of multimorbidity (≥2 comorbidities) was higher in the SOC + NMES group compared with the SOC group (45.2% vs 29.2%), although this difference was not statistically significant (p = 0.27). A similar proportion of participants reported no comorbidities (35.5% vs 33.3%, p = 1.00).

The only statistically significant difference between groups was observed in the endocrine/metabolic category, which was more common in the SOC + NMES group (38.71% vs 12.50%, p = 0.04). No other between-group differences reached statistical significance.

### Surgical Procedures

Table 3 presents the distribution of surgical procedures by treatment group. Participants underwent a range of procedures, with no statistically significant differences observed between the SOC + NMES and SOC groups (Fisher’s exact test).

**Table 3.** Distribution of surgical procedures in the modified intention-to-treat population by treatment group. Values are presented as number (percentage). Between-group comparisons were performed using Fisher’s exact test; no statistically significant differences were observed.

| Procedure | SOC<br>(n=16), n (%) | SOC + NMES (n=26),<br>n (%) | p value |
| --- | --- | --- | --- |
| Hallux valgus correction | 12 (75.0%) | 13 (50.0%) | 0.2 |
| Weil osteotomy | 1 (6.3%) | 1 (3.8%) | 1.00 |
| Lesser toe surgery | 1 (6.3%) | 4 (15.4%) | 0.63 |
| 1st MTPJ cheilectomy | 2 (12.5%) | 3 (11.5%) | 1.00 |
| 1<sup>st</sup> MTPJ fusion | 0 (0%) | 2 (7.7%) | 0.52 |
| Forefoot reconstruction | 0 (0%) | 3 (11.5%) | 0.28 |

### Primary Endpoint

#### Edema

Figure 4 shows the percentage difference in FO8 measurements between the involved and uninvolved limbs at baseline and at day 14. At baseline, both the SOC + NMES and SOC groups demonstrated small, non–statistically significant differences between limbs.

**Figure 4.**
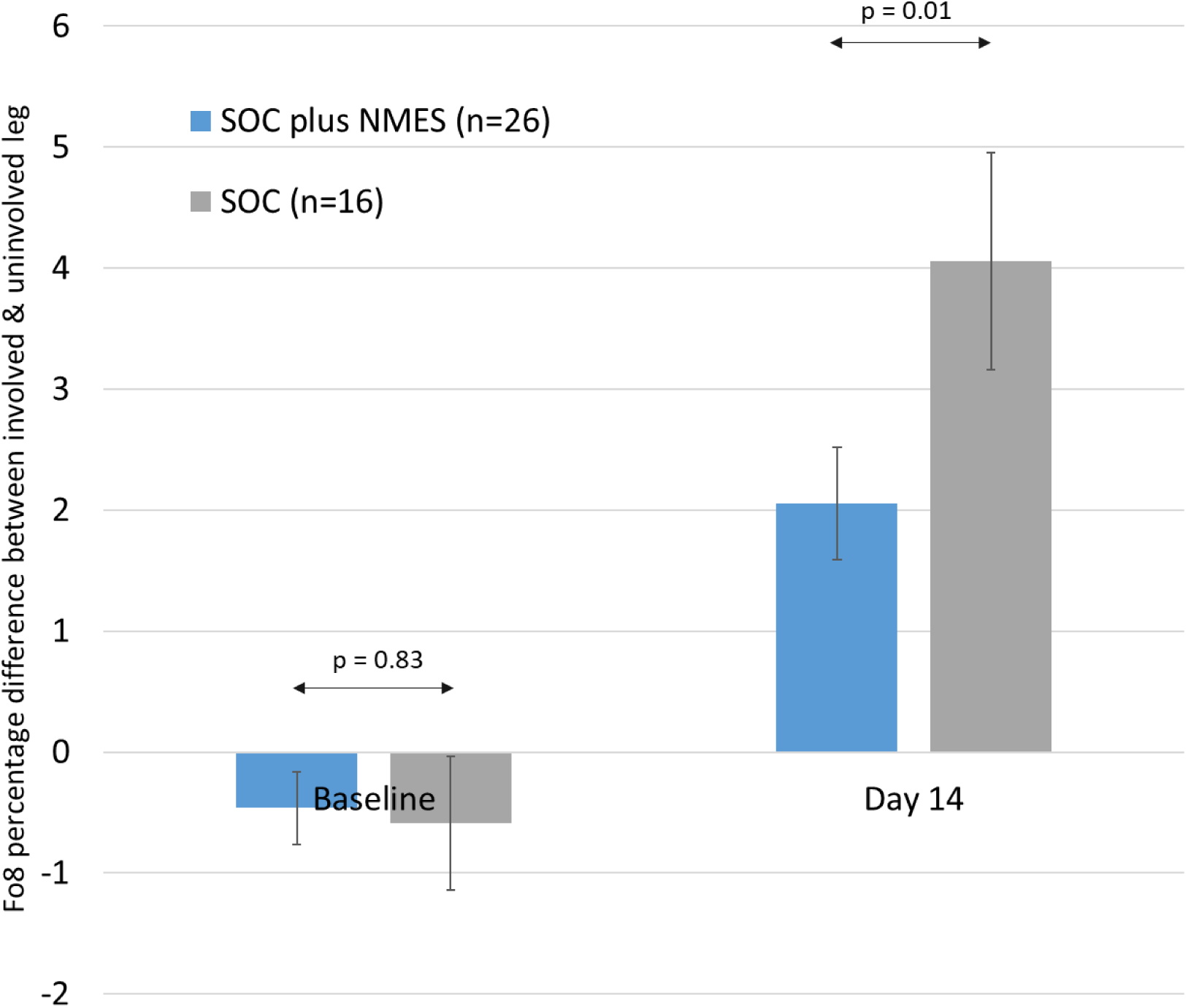
Percentage difference in FO8 measurements between the involved and uninvolved limbs at baseline and day 14 by treatment group. At baseline, no significant difference was observed between groups. At day 14, the SOC group demonstrated greater swelling compared with the SOC + NMES group (4.0% vs 2.0%; p = 0.01). Error bars represent 95% confidence intervals.

At day 14, both groups showed increased swelling in the involved limb compared with the uninvolved limb. The SOC + NMES group demonstrated a mean difference of 2.0% (95% CI 1.6%–2.5%), whereas the SOC group demonstrated a mean difference of 4.0% (95% CI 3.2%–4.9%). This indicates that postoperative swelling was approximately twofold greater in the SOC group compared with the SOC + NMES group. The between-group difference was statistically significant (p = 0.01, unpaired t-test), indicating reduced postoperative edema with NMES.

Figure 5 shows the change in FO8 measurement in the involved limb between baseline and day 14. In the SOC + NMES group, the mean increase was 0.82 cm (95% CI 0.28–1.41 cm), compared with 3.16 cm (95% CI 2.28–3.96 cm) in the SOC group. This corresponds to a 74% reduction in swelling in the SOC + NMES group relative to SOC and was statistically significant (p = 0.02).

**Figure 5.**
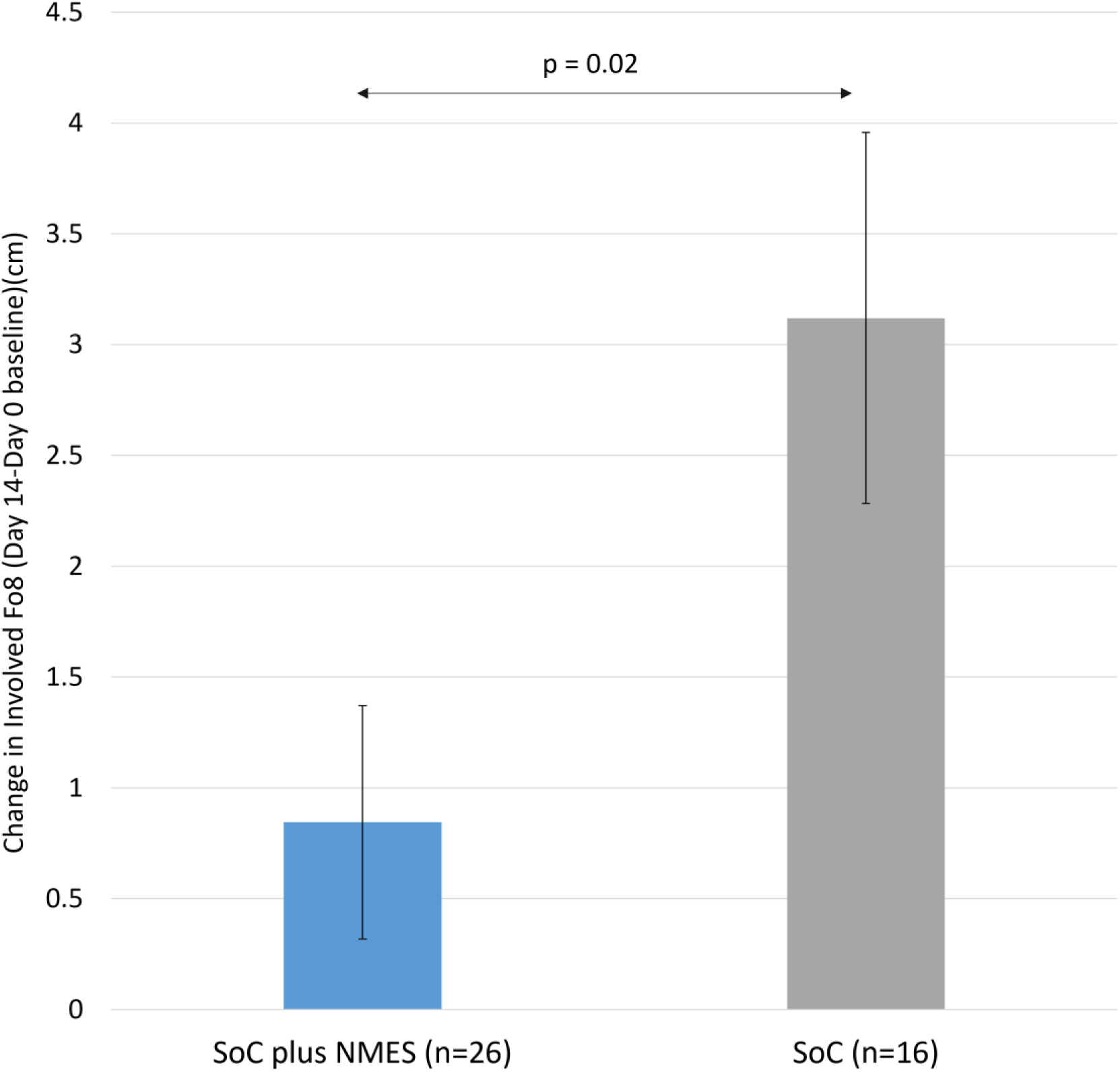
Change in FO8 measurement in the involved limb between baseline and day 14 by treatment group. The SOC group demonstrated a greater increase in swelling compared with the SOC + NMES group (3.16 cm vs 0.82 cm; p = 0.02). Error bars represent 95% confidence intervals.

### Surgical Wound Healing

For this analysis, complete wound healing was defined as full epithelialization of the surgical incision, with no drainage, erythema, dehiscence, or requirement for ongoing dressings at the 14-day postoperative visit. Figure 6 shows the proportion of wounds healed at day 14 in the SOC and SOC + NMES groups. In the SOC + NMES group, 20 of 26 wounds (77%) were healed, compared with 6 of 15 wounds (40%) in the SOC group (data available for 15 participants). This represents an absolute difference of 37%, with healing occurring approximately twice as often in the SOC + NMES group (relative risk 1.93). Despite the small sample size, this difference reached statistical significance (p < 0.05, Fisher’s exact test).

**Figure 6.**
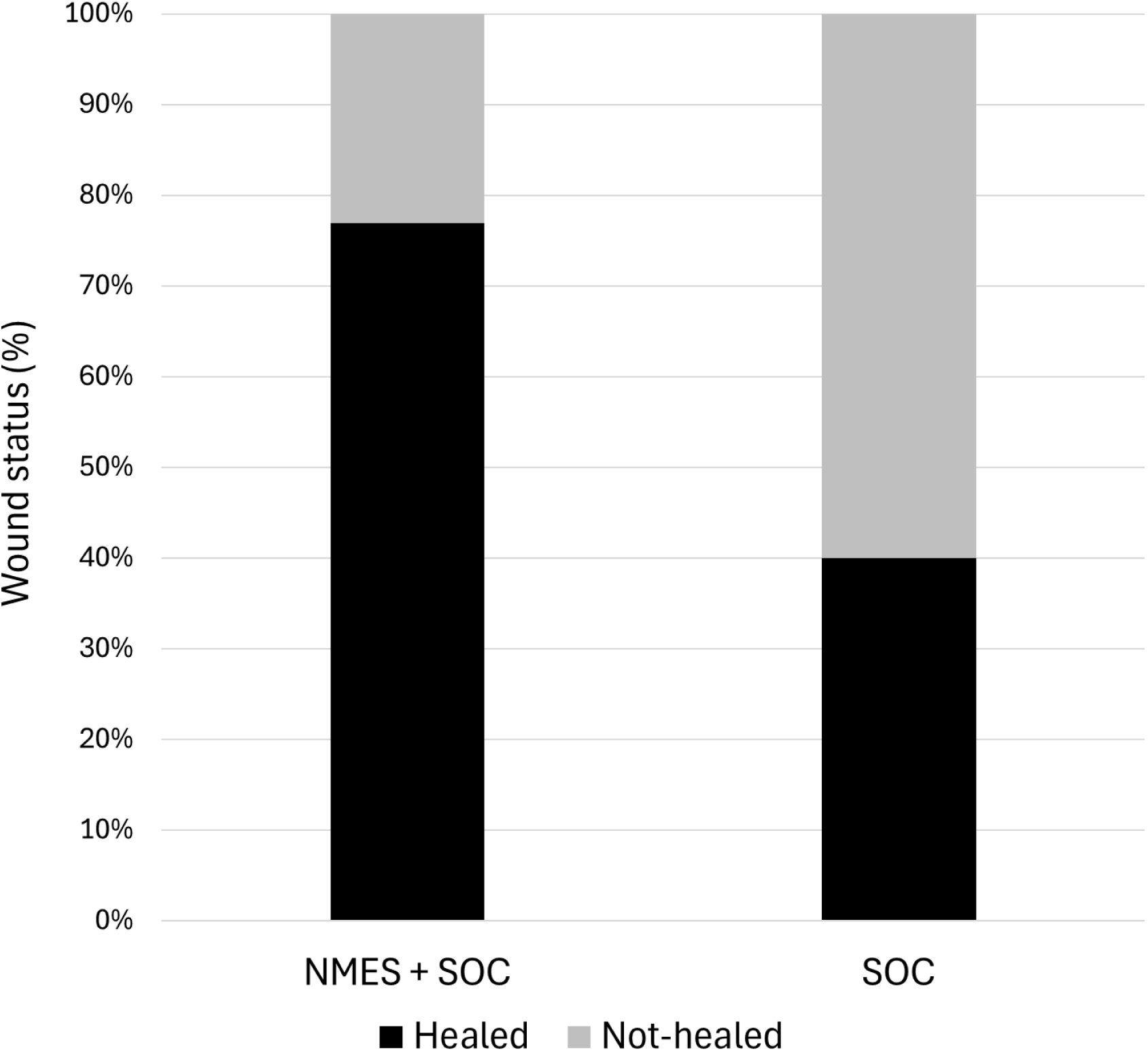
Surgical wound healing status at 14 days postoperatively by treatment group. A greater proportion of wounds were fully healed in the SOC + NMES group compared with the SOC group (77% vs 40%; p < 0.05). Black bars represent healed wounds, and grey bars represent wounds that had not healed at day 14.

### Patient Reported Outcome Measures

Figure 7 presents the results of the Manchester-Oxford Foot Questionnaire (MOXFQ),^24^ which has been shown to be more responsive and reliable in the context of foot and ankle surgery than the Self-Reported Foot and Ankle Score (SEFAS) and other non–foot-specific quality-of-life measures.^25^ For each index, values for the SOC + NMES group are shown in blue (baseline followed by day 14), and values for the SOC group are shown in grey (baseline followed by day 14). Within-group comparisons between baseline and day 14 were performed using paired t-tests. This study was not powered to detect statistically significant between-group differences in patient-reported outcome measures (PROMs), which are inherently less sensitive than objective quantitative measures.

**Figure 7.**
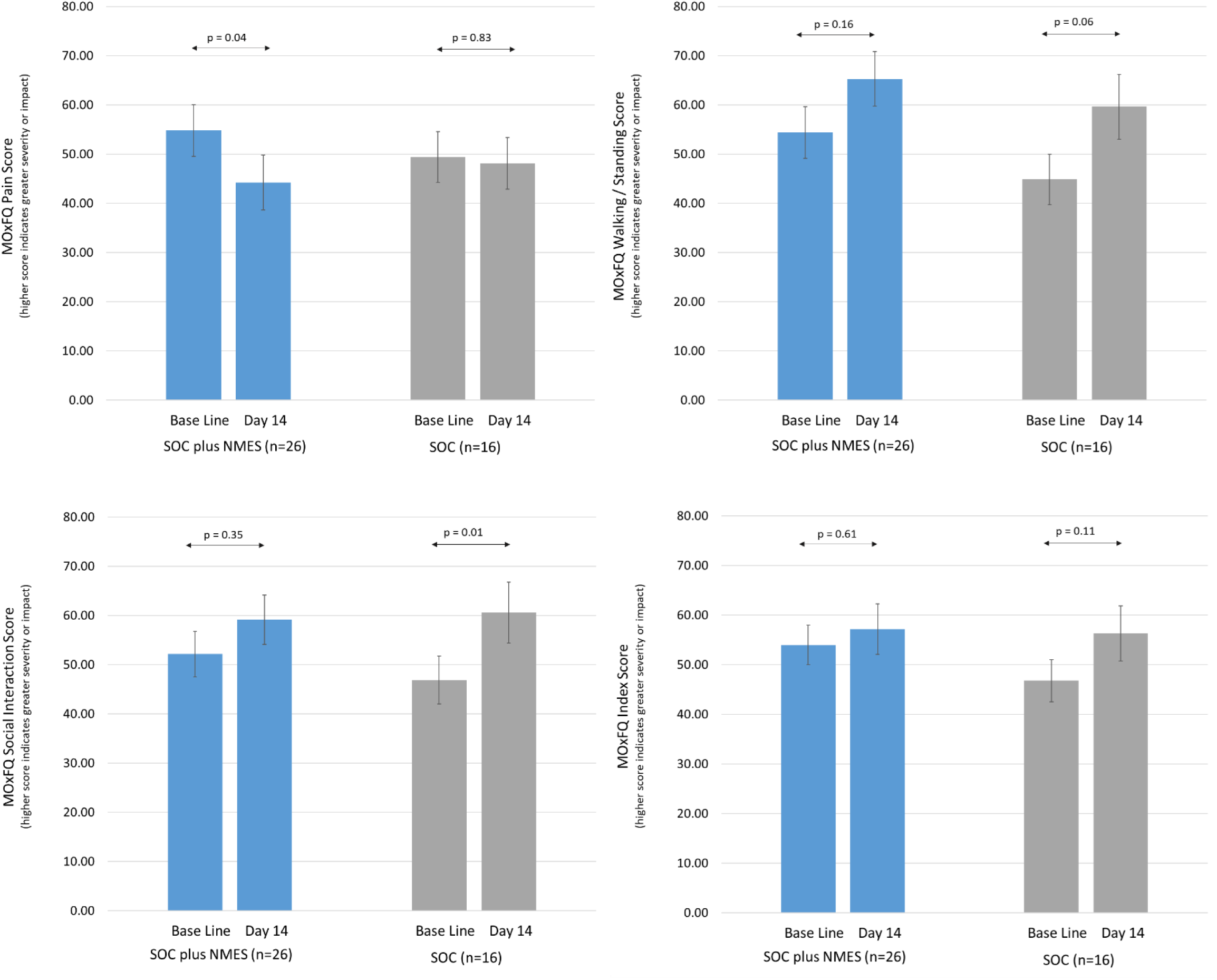
Manchester-Oxford Foot Questionnaire (MOXFQ) scores at baseline and day 14 by treatment group. Scores are shown for (A) pain, (B) walking/standing, (C) social interaction, and (D) overall index. Higher scores indicate greater severity. Within-group comparisons between baseline and day 14 were performed using paired *t*-tests. Error bars represent standard error.

The minimal clinically important change (MCIC) for MOXFQ has been estimated at 13 points for each domain, with minimum detectable change (MDC_90_) values of 11, 12, and 16 for pain, walking, and social interaction scores, respectively.^26^ In the context of hallux valgus surgery, the estimated MCIC values are higher, at 16, 12, and 24 for pain, walking, and social domains, respectively.^27^ These thresholds represent a substantial proportion of the total score range. As shown in Figure 7, none of the observed differences, either between groups or over time, reached the threshold for clinical importance, likely reflecting variability in questionnaire responses.

Although not statistically significant or clinically meaningful, trends across all indices favoured the SOC + NMES group compared with the SOC group. In the pain domain, the SOC + NMES group showed a statistically significant improvement at day 14 (p = 0.03), whereas no change was observed in the SOC group. In the other domains, scores generally increased (indicating worse symptoms) at day 14. This deterioration reached statistical significance in the social interaction domain in the SOC group (p = 0.01) and approached significance for the walking/standing domain (p = 0.06). No statistically significant changes were observed in these domains in the SOC + NMES group.

### Pain

Figure 8 shows pain as measured by the visual analog scale (VAS). Both groups demonstrated significant reductions in pain from baseline to day 14. The SOC + NMES group showed a significant reduction (p = 0.007), as did the SOC group (p = 0.03). Between-group differences were not formally assessed, as the study was not powered to detect differences in this outcome.

**Figure 8.**
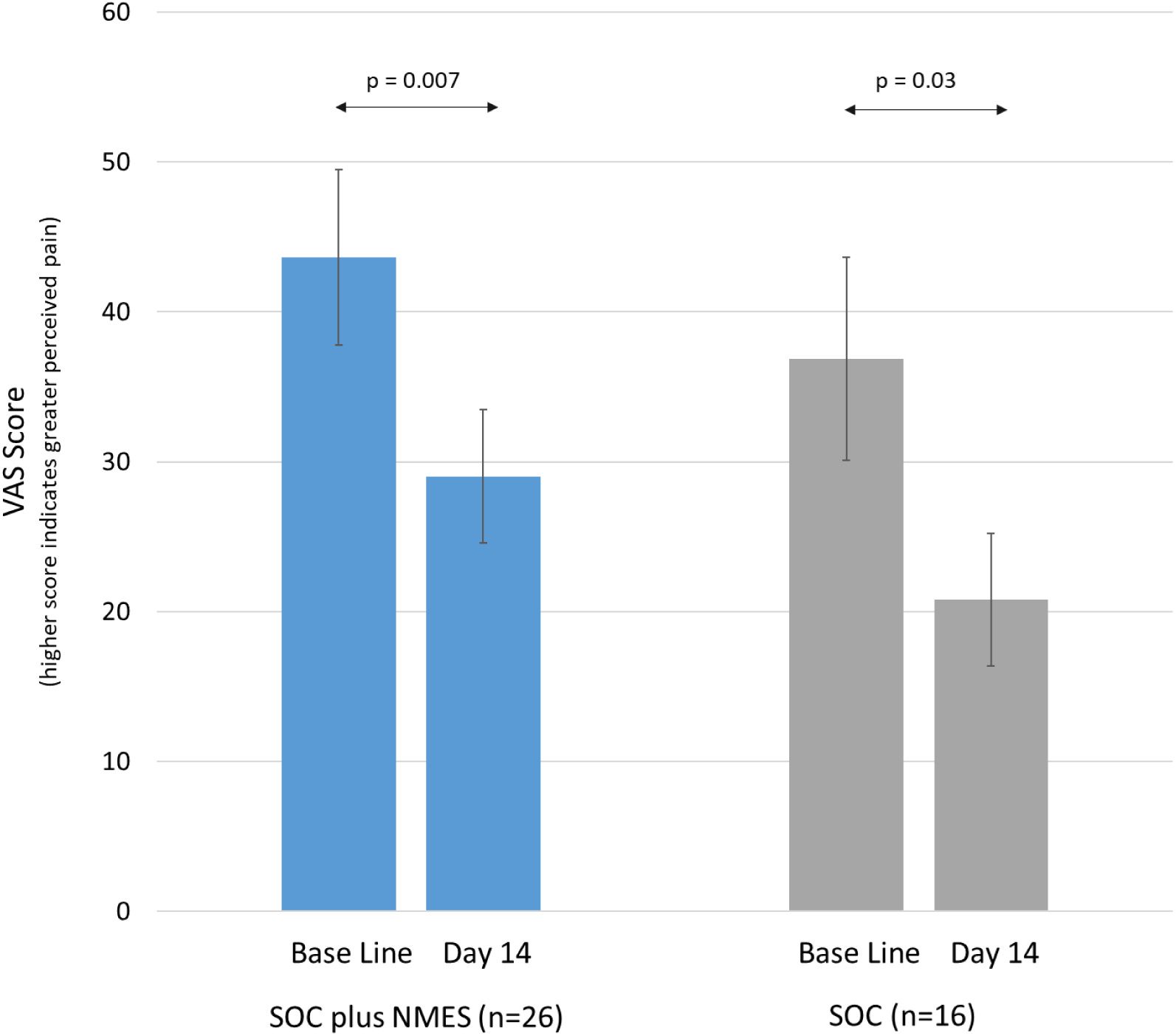
Pain scores measured using the visual analog scale (VAS) at baseline and day 14 by treatment group. Both groups demonstrated significant reductions in pain over time. Error bars represent standard error.

### Adverse Events

Adverse events were categorized as non–device-related, non-serious adverse events (AEs); device-related, non-serious adverse events (ADEs); non– device-related serious adverse events (SAEs); and device-related serious adverse events (SADEs). Table 4 presents the rate of reported adverse events, expressed per 100 patient-days.

**Table 4.** Adverse events per 100 patient days.

| **Event type** | **SOC plus NMES** | **SOC-only** |
| --- | --- | --- |
| AE (non–device-related, non-serious) | 1.16 | 1.43 |
| ADE (device-related, non-serious) | 1.00 | 0.00 |
| SAE (non–device-related, serious) | 0.00 | 0.00 |
| SADE (device-related, serious) | 0.00 | 0.00 |

No device-related adverse events were observed in the SOC group, as no device was used. No serious device-related adverse events were observed in the SOC + NMES group. The small number of device-related adverse events in the SOC + NMES group were limited to skin irritation.

## Discussion

Delayed healing of surgical wounds has been associated with higher complication rates^28^ and poorer outcomes^29^ and may account for up to 20% of healthcare-associated infections.^30^ The annual cost to the NHS of unhealed surgical wounds is estimated at £985 million,^31^ with a single wound costing approximately £141 per additional week of healing.^32^ The use of NMES may reduce wound healing time and thereby lower healthcare costs per patient.

Although the sample size was small for frequency comparison, surgical wounds healed significantly more frequently (p < 0.05) in patients receiving SOC + NMES compared with SOC alone. A plausible explanation for this observation is the increase in microcirculatory blood flow associated with NMES,^33–34^ which is consistent with previous findings demonstrating accelerated healing in chronic wounds.^35–36^ The reduction in venous stasis^37^ with NMES has also been associated with benefits in thromboprophylaxis^38–39^ and may be relevant in the context of recommendations from the NHS Hallux Valgus Think Tank.

Exercise is recognized as beneficial to recovery following bunion surgery,^40–41^ and early mobilization is considered important.^42^ However, return to full activity is often prolonged, frequently exceeding 100 days.^43^ The NMES device offers some of the benefits of a gentle exercise regimen without the need for weight-bearing.^44–45^ By activating the muscle pumps of the lower limb in a manner analogous to walking, NMES enhances fluid clearance^46–47^ without excessive joint movement. This mechanism has also been shown to reduce edema in other clinical settings, including ankle fracture.^48–49^

Patients receiving SOC + NMES demonstrated a 74% reduction in edema compared with SOC alone. Despite the relatively small sample size, this effect was statistically significant (p = 0.02).

The peak of postoperative edema likely occurs before day 14,^50^ and therefore the full temporal dynamics of this effect cannot be determined from the present analysis. It is possible that NMES reduces peak edema, accelerates resolution, or both. Nevertheless, the reduction in edema observed at day 14 may have important implications for recovery time, early mobilization, and overall outcomes.

A previous randomized controlled trial has demonstrated a significant reduction in postoperative swelling with NMES in patients undergoing foot and ankle surgery. This sub-analysis indicates that this effect is also present in patients undergoing forefoot surgery and is accompanied by improved wound healing at day 14.

## Limitations

This study has several limitations. The heterogeneity of the patient population makes precise matching between groups challenging, although this was partially mitigated by using contralateral limb comparison and by assessing changes relative to baseline measurements. In addition, the reduced sample size in this sub-analysis, compared with that originally calculated to power the parent study, limits statistical power.

The open-label design and lack of blinding may have introduced bias, particularly in subjective outcome measures. Outcome assessors were not blinded to treatment allocation, and FO8 measurements were performed by multiple trained assessors across sites. Although standardized training, use of a predefined measurement protocol, repeated measurements with averaging, and consistent equipment were employed to minimize variability, some measurement variability may remain. Any residual variability would be expected to affect both groups similarly. Furthermore, outcome assessment at a single postoperative time point (day 14) limits understanding of the full temporal profile of edema and recovery. As this represents a post hoc subgroup analysis, the findings should be interpreted with caution and considered hypothesis-generating.

## Acknowledgement

We gratefully acknowledge the involvement of Maidstone and Tunbridge Wells NHS Trust, Royal National Orthopaedic Hospital NHS Trust, University Hospital Vall d’Hebron, Hospital Clínic Barcelona, and OrthoNorCal, Inc. in supporting the conduct of the study.

## Data Availability Statement

The data supporting the findings of this study are available from the corresponding author upon reasonable request, subject to applicable data protection and confidentiality requirements.

